# The recurrent deep intronic pseudoexon-inducing variant *COL6A1* c.930+189C>T results in a consistently severe phenotype of COL6-related dystrophy: Towards clinical trial readiness for splice-modulating therapy

**DOI:** 10.1101/2024.03.29.24304673

**Authors:** A. Reghan Foley, Véronique Bolduc, Fady Guirguis, Sandra Donkervoort, Ying Hu, Rotem Orbach, Riley M. McCarty, Apurva Sarathy, Gina Norato, Beryl B. Cummings, Monkol Lek, Anna Sarkozy, Russell J. Butterfield, Janbernd Kirschner, Andrés Nascimento, Daniel Natera-de Benito, Susana Quijano-Roy, Tanya Stojkovic, Luciano Merlini, Giacomo Comi, Monique Ryan, Denise McDonald, Pinki Munot, Grace Yoon, Edward Leung, Erika Finanger, Meganne E. Leach, James Collins, Cuixia Tian, Payam Mohassel, Sarah B. Neuhaus, Dimah Saade, Benjamin T. Cocanougher, Mary-Lynn Chu, Mena Scavina, Carla Grosmann, Randal Richardson, Brian D. Kossak, Sidney M. Gospe, Vikram Bhise, Gita Taurina, Baiba Lace, Monica Troncoso, Mordechai Shohat, Adel Shalata, Sophelia H.S. Chan, Manu Jokela, Johanna Palmio, Göknur Haliloğlu, Cristina Jou, Corine Gartioux, Herimela Solomon-Degefa, Carolin D. Freiburg, Alvise Schiavinato, Haiyan Zhou, Sara Aguti, Yoram Nevo, Ichizo Nishino, Cecilia Jimenez-Mallebrera, Shireen R. Lamandé, Valérie Allamand, Francesca Gualandi, Alessandra Ferlini, Daniel G. MacArthur, Steve D. Wilton, Raimund Wagener, Enrico Bertini, Francesco Muntoni, Carsten G. Bönnemann

**Affiliations:** Neuromuscular and Neurogenetic Disorders of Childhood Section, Neurogenetics Branch, National Institute of Neurological Disorders and Stroke, NIH, Bethesda, MD 20892, USA; Dana-Dwek Children’s Hospital, Tel Aviv 64239, Israel; Clinical Trials Unit, National Institute of Neurological Disorders and Stroke, NIH, Bethesda, MD 20892, USA; Broad Institute of MIT and Harvard, Cambridge, MA 02142, USA; Dubowitz Neuromuscular Centre, UCL Great Ormond Street Institute of Child Health and Great Ormond Street Hospital for Children, London WC1N 1EH, UK; Departments of Neurology and Pediatrics, University of Utah, Salt Lake City, UT 84132, USA; Department of Neuropediatrics and Muscle Disorders, Medical Center – University of Freiburg, Faculty of Medicine, Freiburg 79110, Germany; Neuromuscular Unit, Neuropediatrics Department, Institut de Recerca Sant Joan de Déu, Hospital Sant Joan de Déu. CIBERER ISCIII. Barcelona 08950, Spain; Garches Neuromuscular Reference Center, Child Neurology and ICU Department, APHP Raymond Poincare University Hospital (UVSQ Paris Saclay), Garches 92380, France; Centre de Référence des Maladies Neuromusculaires Nord/Est/Île-de-France, Institut de Myologie, Hôpital Pitié-Salpêtrière, AP-HP, Paris 75013, France; Department of Biomedical and Neuromotor Sciences, University of Bologna, Bologna 40126, Italy; Neurology Unit, Fondazione IRCCS Ca’ Granda Ospedale Maggiore Policlinico, Milan, Italy; Department of Neurology, The Royal Children’s Hospital, Parkville, VIC 3052, Australia; Department of Neurodisability, Children’s Health Ireland at Tallaght, Dublin 24 Ireland; Department of Pediatrics, Division of Clinical and Metabolic Genetics, The Hospital for Sick Children, University of Toronto, Toronto, ON M5G 1X8, Canada; Department of Pediatrics and Child Health, University of Manitoba, Winnipeg, MB R3A 1S1, Canada; Department of Pediatrics and Neurology, Oregon Health & Science University, Portland, OR 97239, USA; Divisions of Neurology and Pediatrics, Cincinnati Children’s Hospital Medical Center, Cincinnati, OH 45229, USA; Division of Medical Genetics, Department of Pediatrics, Duke University, Durham, NC 27710, USA; Department of Neurology, New York University School of Medicine, New York, NY 10016, USA; Division of Neurology, Nemours Children’s Hospital Delaware, Wilmington, DE 19803, USA; Department of Neurology, Rady Children’s Hospital University of California San Diego, San Diego, CA 92123, USA; Department of Neurology, Gillette Children’s Specialty Healthcare, St Paul, MN 55101, USA; Department of Neurology, Dartmouth Hitchcock Medical Center, Lebanon, NH 03766, USA; Department of Neurology and Pediatrics, University of Washington, Seattle, WA 98105, USA; Departments of Pediatrics and Neurology, Rutgers Robert Wood Johnson Medical School, Rutgers University, New Brunswick, NJ 08901, USA; Children’s Clinical University Hospital, Medical Genetics and Prenatal Diagnostic Clinic, Riga 1004, Latvia; Riga East Clinical University, Institute of Clinical and Preventive Medicine of the University of Latvia, Riga 1586, Latvia; Pediatric Neuropsychiatry Service, Hospital Clínico San Borja Arriarán, Pediatric Department, Universidad de Chile, Santiago 1234, Chile; The Genomics Unit, Sheba Cancer Research Center, Sheba Medical Center, Ramat Gan 52621, Israel; The Simon Winter Institute for Human Genetics, Bnai Zion Medical Center, The Ruth and Bruce Rappaport Faculty of Medicine, Technion-Israel Institute of Technology, Haifa 32000, Israel; Department of Paediatrics and Adolescent Medicine, Li Ka Shing Faculty of Medicine, The University of Hong Kong, Hong Kong, Special Administrative Region, China; Clinical Neurosciences, University of Turku, Turku, Finland and Neurocenter, Turku University Hospital, Turku 20520, Finland; Neuromuscular Research Center, Tampere University and Tampere University Hospital, Tampere 33101, Finland; Division of Pediatric Neurology, Department of Pediatrics, Hacettepe University Faculty of Medicine, Ankara 06230, Turkey; Pathology department, Institut de Recerca Sant Joan de Déu, Hospital Sant Joan de Déu, Barcelona 08950, Spain; INSERM, Institut de Myologie, Centre de Recherche en Myologie, Sorbonne Université, Paris 75013, France; Center for Biochemistry, Medical Faculty, University of Cologne, Cologne 50931, Germany; National Institute of Health Research, Great Ormond Street Hospital Biomedical Research Centre, Genetics and Genomic Medicine Research and Teaching Department, Great Ormond Street Institute of Child Health, University College London, London WC1N 1EH, UK; Neurodegenerative Disease Department, UCL Queen Square Institute of Neurology, University College London, London WC1N 3BG, UK; Institute of Pediatric Neurology, Schneider Children’s Medical Center of Israel, Petach Tikva, Israel, Sackler Faculty of Medicine, Tel Aviv University, Tel Aviv 69978, Israel; Department of Neuromuscular Research, National Institute of Neuroscience, National Center of Neurology and Psychiatry, Tokyo 187-8502, Japan; Laboratorio de Investigación Aplicada en Enfermedades Neuromusculares, Unidad de Patología Neuromuscular, Servicio de Neuropediatría, Institut de Recerca Sant Joan de Déu, Barcelona 08950, Spain; Department of Paediatrics, University of Melbourne, The Murdoch Children’s Research Institute, Parkville, VIC 3052, Australia; Unit of Medical Genetics, Department of Medical Sciences and Department of Mother and Child, University Hospital S. Anna Ferrara, Ferrara 44121, Italy; Centre for Molecular Medicine and Innovative Therapeutics, Health Futures Institute, Murdoch University; Centre for Neuromuscular and Neurological Disorders, Perron Institute for Neurological and Translational Science, The University of Western Australia, Nedlands, WA 6009, Australia; Research Unit of Neuromuscular and Neurodegenerative Disorders, IRCCS Ospedale Pediatrico Bambino Gesù, Rome 00146, Italy; National Institute of Health Research, Great Ormond Street Hospital Biomedical Research Centre, London WC1N 1EH, UK

## Abstract

Collagen VI-related dystrophies (COL6-RDs) manifest with a spectrum of clinical phenotypes, ranging from Ullrich congenital muscular dystrophy (UCMD), presenting with prominent congenital symptoms and characterised by progressive muscle weakness, joint contractures and respiratory insufficiency, to Bethlem muscular dystrophy, with milder symptoms typically recognised later and at times resembling a limb girdle muscular dystrophy, and intermediate phenotypes falling between UCMD and Bethlem muscular dystrophy. Despite clinical and immunohistochemical features highly suggestive of COL6-RD, some patients had remained without an identified causative variant in *COL6A1*, *COL6A2* or *COL6A3*. With combined muscle RNA-sequencing and whole-genome sequencing we uncovered a recurrent, *de novo* deep intronic variant in intron 11 of *COL6A1* (c.930+189C>T) that leads to a dominantly acting in-frame pseudoexon insertion. We subsequently identified and have characterised an international cohort of forty-four patients with this *COL6A1* intron 11 causative variant, one of the most common recurrent causative variants in the collagen VI genes. Patients manifest a consistently severe phenotype characterised by a paucity of early symptoms followed by an accelerated progression to a severe form of UCMD, except for one patient with somatic mosaicism for this *COL6A1* intron 11 variant who manifests a milder phenotype consistent with Bethlem muscular dystrophy. Characterisation of this individual provides a robust validation for the development of our pseudoexon skipping therapy. We have previously shown that splice-modulating antisense oligomers applied *in vitro* effectively decreased the abundance of the mutant pseudoexon-containing COL6A1 transcripts to levels comparable to the *in vivo* scenario of the somatic mosaicism shown here, indicating that this therapeutic approach carries significant translational promise for ameliorating the severe form of UCMD caused by this common recurrent *COL6A1* causative variant to a Bethlem muscular dystrophy phenotype.

## Introduction

Ullrich congenital muscular dystrophy (UCMD [MIM 254090]), Bethlem myopathy or rather muscular dystrophy (BM [MIM 158810]) and intermediate phenotypes form a subgroup within the congenital muscular dystrophies (CMDs) known as the collagen VI-related dystrophies (COL6-RDs)^1–3^ and result from recessively or dominantly acting causative variants in any of the three collagen VI genes (*COL6A1*, *COL6A2* or *COL6A3*).^4,5^ Ullrich congenital muscular dystrophy was first described in 1930 by Dr. Otto Ullrich, who termed the condition ‘Skleratonische Muskeldystrophie’ (scleratonic muscular dystrophy), noting evidence of congenital weakness associated with proximal joint contractures and distal joint laxity.^6,7^ The first signs of UCMD can manifest *in utero* with decreased fetal movement frequently reported.^1,6–9^ At birth, UCMD patients classically demonstrate hypotonia, proximal joint contractures, hip dislocation(s), prominent calcanei and distal joint hyperlaxity, typically resulting in abnormal positioning of the hands and feet (with hands in a position of wrist flexion, resting against the ventral surface of the forearms and feet in a position of dorsiflexion, resting against the anterior surface of the lower leg). Torticollis and kyphoscoliosis can often be seen at birth, as well. While children with UCMD may achieve independent ambulation, this ability is lost in early childhood^1,9–11^ typically by 10 years of age.^3,12^ After becoming wheelchair-dependent, most UCMD patients demonstrate muscle weakness which appears to be relatively stable (as far as can be assessed within restricted range of movement) while joint contractures continue to progress, compounding the motor limitations resulting from the muscle weakness and thus significantly contributing to the overall level of disability.^13^ While some UCMD patients demonstrate spinal stiffness without an evident spinal curvature, the vast majority develop progressive scoliosis which can appear as early as the preschool years or even congenitally.^3,12^ An early onset of an invariable decline in respiratory function with early hypoventilation is a salient clinical feature of UCMD, necessitating the initiation of nocturnal non-invasive ventilation by an average age of 11 years.^2,14^

Bethlem myopathy (BM) was first described in 1976 by Drs. Jaap Bethlem and George K. van Wijngaarden and is characterised by slowly progressive muscle weakness and distally pronounced joint contractures.^15^ While often described as a slowly progressive ‘myopathy’ of adulthood, muscle histology progresses over time to include dystrophic findings, and thus the term ‘Bethlem muscular dystrophy’ more accurately describes this condition.^16^ The categorisation of Bethlem muscular dystrophy (MD) within the congenital muscular dystrophies likely relates, in part, to the fact that symptoms of Bethlem muscular dystrophy can present as early as birth. In particular, hypotonia, neck flexion weakness, torticollis and joint contractures are among the early symptoms reported. Progressive contractures of the Achilles tendons and elbows usually manifest by the end of the first decade. Patients with Bethlem MD develop proximal muscle weakness but typically maintain the ability to ambulate into adulthood. By 50 years of age, however, more than 2/3 of patients rely on the use of a wheelchair to aide ambulation, classically for outdoor use while independent ambulation indoors is usually maintained.^17^ While some adults with Bethlem muscular dystrophy develop nocturnal hypoventilation, this does not occur uniformly, as seen in a large natural history study of pulmonary function in the COL6-RDs which did not demonstrate a statistically significant correlation between age and decrease in pulmonary function in patients with Bethlem MD.^2^

Ullrich congenital muscular dystrophy and Bethlem muscular dystrophy were initially viewed as two separate phenotypic entities; however, subsequently it became apparent that there is a continuous spectrum of phenotypes, with UCMD at the severe end of the spectrum, Bethlem MD along the mild end, and so-called ‘intermediate’ phenotypes in between the classical UCMD and Bethlem MD phenotypes.^2,3,13,18^ A large-scale international natural history study of COL6-RD patients helped in elucidating the parameters of intermediate COL6-RD. Patients in this group uniformly achieved ambulation and walked longer than UCMD patients but never achieved the ability to jump or run, in contrast to patients with Bethlem MD. For this intermediate COL6-RD group, loss of ambulation occurred by approximately 19 years of age, and nocturnal non-invasive ventilation (NIV) was needed by approximately 21.5 years of age.^2^

Anticipatory care of the uniformly progressive decline in respiratory function which characterises UCMD and intermediate COL6-RD, including the timely initiation of non-invasive ventilation, is essential for decreasing morbidity and mortality, and this care relies on the clinical recognition of COL6-RD. Despite clinical and immunohistochemical features highly suggestive of COL6-RD, some patients have remained without an identified causative variant in the *COL6A1*-*3* genes. By using a combination of muscle RNA-seq and whole-genome sequencing in four such patients, we uncovered a recurrent, *de-novo* deep-intronic variant in intron 11 of *COL6A1* (c.930+189C>T) that leads to a dominantly acting in-frame pseudoexon insertion.^19,20^ Targeted Sanger sequencing for this variant in intron 11 of *COL6A1* to complement panel and whole exome testing has revealed that this deep intronic *de-novo* causative variant is a surprisingly common cause of UCMD. Here we describe the consistent phenotype associated with this *COL6A1* intron 11 causative variant, which manifests with a paucity of symptoms congenitally and an accelerated progression to a phenotype of UCMD, except in one patient with somatic mosaicism for this *COL6A1* variant who manifests a significantly milder phenotype which is consistent with Bethlem muscular dystrophy.

## Materials and Methods

### Study subjects

Patients were identified through their local neurology clinics. Written informed consent and age-appropriate assent for research studies, procedures, and clinical photographs was obtained by a qualified investigator. Ethical approval was obtained via the Institutional Review Board of the National Institutes of Health (protocol 12-N-0095), the University College London Research Ethics Committee (13/LO/1894) and the MRC Centre for Neuromuscular Disease Biobank London Research Ethics Committee (06/Q0406/33). Medical history was obtained, and clinical evaluation, muscle imaging and biopsies, were performed as part of the standard neurologic evaluation. Muscle MRI was performed using conventional T1-weighted spin echo of the lower extremities. Muscle ultrasound images were obtained using a Siemens/Acuson S2000 with an 18 MHz linear probe. Blood, skin biopsies, muscle biopsies and urine samples were obtained according to standard procedures. Saliva samples were collected using the oragene-discover kit (DNA Genotek, Ottawa, Canada).

### Biospecimen processing

Fibroblasts were cultured from fresh skin biopsies using a standard enzymatic digestion methodology. DNA was extracted from blood, skin fibroblasts, saliva and the pelleted fraction of the urine specimen using the Gentra Puregene Blood kit (Qiagen, Germantown, MD). RNA was obtained from blood following the manual purification of total RNA for human whole blood collected in PAXgene blood RNA tubes protocol (PreAnalytiX/Qiagen, Germantown, MD). RNA was obtained from skin fibroblasts using Trizol (ThermoFisher Scientific, Waltham, MA). For fresh skin biopsy samples, specimens were frozen in liquid nitrogen, pulverized using a mortar and pillar and homogenized in Trizol. DNA and RNA were both obtained using the interphase/organic and the aqueous phase of Trizol, respectively, following manufacturer’s instructions.

### Sanger sequencing

Endpoint PCR was performed on gDNA samples using Takara LA Taq (Takara Bio Inc., Shiga, Japan) per manufacturer’s specifications. Sequences of the primers were as follow: 5’-TGTTGGGTACCAGGGAATGAAGGT-3’, 5’-AAACGAAGGCAGGAGTCAGA-3’. PCR products were sent for Sanger sequencing to Genewiz (Azenta Life Sciences, South Plainfield, NJ).

### Pseudoexon amplification and quantification

To quantify the degree of mosaicism, a Taqman assay was designed and synthesized by ThermoFisher Scientific as a Custom Taqman SNP Genotyping Assay, in which the probe hybridizing to the reference (C) allele was labeled with VIC, and the probe hybridizing to the variant (T) allele was labeled with FAM. gDNA samples (between 50 and 600 ng of DNA, depending on the tissue source) were amplified on the Bio-Rad QX200 ddPCR system (Bio-Rad, Hercules, CA) at the NCI CCR Genomics Core, using the standard protocol provided by the manufacturer and the assay described above. The fractional abundance of the variant (T) allele over the reference (C) allele was calculated from the determined concentrations (copies/µL).

For expression studies, RNA was converted to cDNA using the SuperScript IV Reverse Transcriptase (ThermoFisher). For end-point PCR and gel electrophoresis, cDNA was amplified using the Kapa HiFi HotStart (Roche Sequencing, Indianapolis, IN), with the following primers: 5’-acctgttgggtaccagggaatgaa-3’, and 5’-accagggtctcctcttggtc-3’. For quantitative PCR, cDNA was amplified using the FastStart Universal Master Mix (Roche Life Science), on the QuantStudio 6 Real-Time PCR instrument (ThermoFisher Scientific). Expression levels of the *COL6A1* transcripts were determined with quantitative PCR assays detecting either transcripts with the pseudoexon (primer 1: 5’-taccagggaatgaagggag-3’, primer 2: 5’-cctggagccctttgctg-3’, probe: 5’-atctggaaggacaaggacagccac-3’) or total *COL6A1* transcripts (primer 1: 5’-ccgactgcgctatcaagaa-3’, primer 2: 5’-aatcaggtacttattctccttcaggt-3’, probe: ROCHE UPL probe #17, Millipore Sigma, Burlington, MA - product now discontinued). Expression of the *COL6A1* with pseudoexon transcripts were normalised to the total *COL6A1* expression. The patient’s samples were calibrated with the control (or average of control) samples, following the 2^-ΔΔCt^ method.

### Muscle immunofluorescence

Frozen muscle tissues were cross-sectioned (10 µm) from control and patient muscle biopsies, fixed in pre-cooled 100% methanol at −20°C for 5 minutes and washed in phosphate-buffered saline (PBS). Sections were blocked in PBS with 10% fetal bovine serum (FBS) and 0.1% Triton X-100 for 30 minutes. Primary antibodies were diluted in the blocking buffer and incubated overnight at 4°C [anti-collagen VI mouse monoclonal antibody MAB3303 (1:2500) and anti-laminin rabbit antibody L9393 (1:800)]. Alexa-568 and Alexa-488-conjugated secondary antibodies (1:500; Invitrogen) were used, and immunofluorescence images were obtained using Zeiss Airy confocal microscope.

### Statistical Analysis

Summary statistics for onset of ambulation, loss of ambulation and onset of non-invasive ventilation were described using mean ± standard deviation. All statistical tests were conducted with a significance level of 0.05. A Mann-Whitney U test was performed to compare the onset of ambulation, loss of ambulation and onset of non-invasive ventilation for UCMD patients heterozygous for the *COL6A1* c.930+189C>T variant to UCMD patients with causative variants in the *COL6* genes (other than the *COL6A1* c.930+189C>T variant). Log-rank tests were performed for the Kaplan-Meier curves.

## Results

### *COL6A1* variant

The *COL6A1* c.930+189C>T variant was identified via targeted Sanger sequencing (N=27),^20^ performed in our laboratory on DNA samples for patients with a clinical phenotype of COL6-RD who remained without an identified causative variant in the COL6 genes (*COL6A1*, *COL6A2* and *COL6A3*) on panel or whole exome sequencing, or so-called ‘causative variant negative’ COL6-RD, and following the initial identification of this variant via RNA sequencing performed in muscle biopsy samples from patients with COL6-RD who were ‘causative variant negative’ (N=4).^19^ Following the inclusion of this variant on diagnostic next generation sequencing panels, it was identified in further patients (N=13), with one of these patients with evidence of somatic mosaicism for the *COL6A1* c.930+189C>T variant.

### Clinical presentation

Forty-four patients were found to harbour the *COL6A1* c.930+189C>T causative variant; one patient (US16) has somatic mosaicism for this variant. In all patients in whom parental segregation testing was performed, the variant was confirmed to be *de novo*. Demographic and clinical features are listed in Table 1. Sixteen patients were identified in the United States (U1-U16), five in the United Kingdom (UK1-5), five in Italy (I1-I5), three in Canada (CA1-CA3), two patients each in France (F1-F2), Spain (S1-S2) and Latvia (L1-L2), and one patient each was identified in Australia (A1), Chile (CH1), Germany (G1), Hong Kong (HK1), Ireland (I1), Israel (IS1), Romania (R1), Turkey (T1) and Finland (FI1). Twenty-seven of the patients are female, and seventeen are male. Three patients were included in a report of a cohort of patients with COL6-RD in Italy.^21^ Phenotypic data on the clinical presentation of patients with a phenotype of UCMD due to causative variants in the *COL6* genes other than the *COL6A1* c.930+189C>T variant were evaluated to serve as a comparison cohort to the *COL6A1* c.930+189C>T specific cohort. Seventeen patients with UCMD, 12 females and 5 males evaluated at the National Institutes of Health (NIH) were included in this comparison cohort (Supplementary Table 1).

**Table 1:** Core phenotypic features of patients with Ullrich congenital muscular dystrophy due to *COL6A1* (c.930+189C>T)

| Patient Identifier | Sex | Race/Ethnicity | Decreased fetal movements | Hip dislocation or hip dysplasia at birth | Hypotonia at birth | Abnormal position of hands/feet at birth | Torticollis at birth |
| --- | --- | --- | --- | --- | --- | --- | --- |
| US1 | F | Hispanic/Mexican | Y | N | N | N | N |
| US2 | M | Caucasian/Ashkenazi Jewish | N | N | N | N | N |
| US3 | M | Hispanic | unknown | N | Y | unknown | N |
| US4 | F | Caucasian/American | Y | N | N | N | N |
| US5 | F | Caucasian/American | Y | Y | Y | N | N |
| US6 | F | Caucasian/American | N | N | N | N | N |
| US7 | F | Caucasian/American | Y | Y | N | N | N |
| US8 | F | Hispanic | N | Y | N | N | N |
| US9† | M | Black/American | N | N | N | N | N |
| US10 | M | Hispanic/Guatemalan and Costa Rican | N | N | N | N | N |
| US11 | M | Hispanic | Y | N | N | Y | Y |
| US12 | M | Asian/Indian | Y | N | Y | N | N |
| US13 | F | Caucasian/American | N | Y | N | unknown | N |
| US14 | F | Caucasian/American | N | N | N | N | N |
| US15 | F | Hispanic/Colombian | Y | Y | N | N | N |
| UK1 | M | Caucasian/British | Y | N | Y | N | Y |
| UK2 | M | Caucasian/Irish | unknown | unknown | unknown | unknown | unknown |
| UK3 | F | Caucasian/British | Y | Y | Y | N | N |
| UK4 | F | Caucasian/Greek | unknown | unknown | unknown | unknown | unknown |
| UK5 | F | Caucasian/Spanish | N | N | N | N | N |
| IT1 | F | Caucasian/Italian | N | Y | N | N | Y |
| IT2 | F | Caucasian/Italian | Y | N | Y | N | N |
| IT3 | M | Caucasian/Italian | Y | N | N | N | N |
| IT4 | F | Caucasian/Italian | N | Y | N | N | Y |
| IT5 | M | Caucasian/Italian | N | N | N | N | N |
| CA1 | F | Asian/Indian | N | N | N | N | N |
| CA2 | M | Asian/Bengali | N | N | N | N | N |
| CA3†† | M | Caucasian/Canadian | N | N | N | N | N |
| F1 | M | Caucasian/French | N | N | Y | Y | N |
| F2 | F | Caucasian/French<br>and African/Algerian | N | Y | N | N | N |
| S1 | M | Caucasian/Spanish | N | N | Y | N | N |
| S2 | F | Caucasian/Spanish | N | N | N | N | N |
| L1 | M | Caucasian/Latvian | N | N | N | Y | N |
| L2 | F | Caucasian/Latvian | Y | Y | Y | N | N |
| A1 | F | Caucasian/Australian | Y | Y | N | N | N |
| CH1 | F | Hispanic/Chilean | Y | Y | N | N | N |
| FI1 | F | Caucasian/Finnish | N | N | N | Y | N |
| G1 | M | Caucasian/German | Y | Y | Y | N | N |
| HK1 | F | Asian/Chinese | N | Y | Y | N | N |
| IR1 | M | Caucasian/Irish | N | N | N | N | N |
| IS1 | F | Caucasian/Arab | N | N | N | N | N |
| R1 | F | Caucasian/Romanian | Y | N | N | Y | N |
| T1 | F | Caucasian/Turkish | Y | N | N | Y | N |
| US16* | F | Caucasian/American | N | N | Y | N | N |

†Died (due to probable *cor pulmonale* related to respiratory insufficiency)
††Died (due to respiratory failure and failure to thrive)
\*This patient has somatic mosaicism for *COL6A1* (c.930+189C>T)
NA = not available

### Phenotype of patients heterozygous for the *COL6A1* c.930+189C>T variant

A history of decreased fetal movements during pregnancy was reported by the respective mothers of 43% (17/40) of the patients. Of those patients with findings documented at birth, 34% (14/41) had hip dislocation and/or hip dysplasia, 27% (11/41) had evidence of hypotonia, 15% (6/39) had abnormal positioning of hands and feet, and 10% (4/41) had torticollis (Fig. 1A). The mean onset of independent ambulation was 1.4 ± 0.3 years, with two patients reported to have never attained independent ambulation (Fig. 1B).

**Figure 1:**
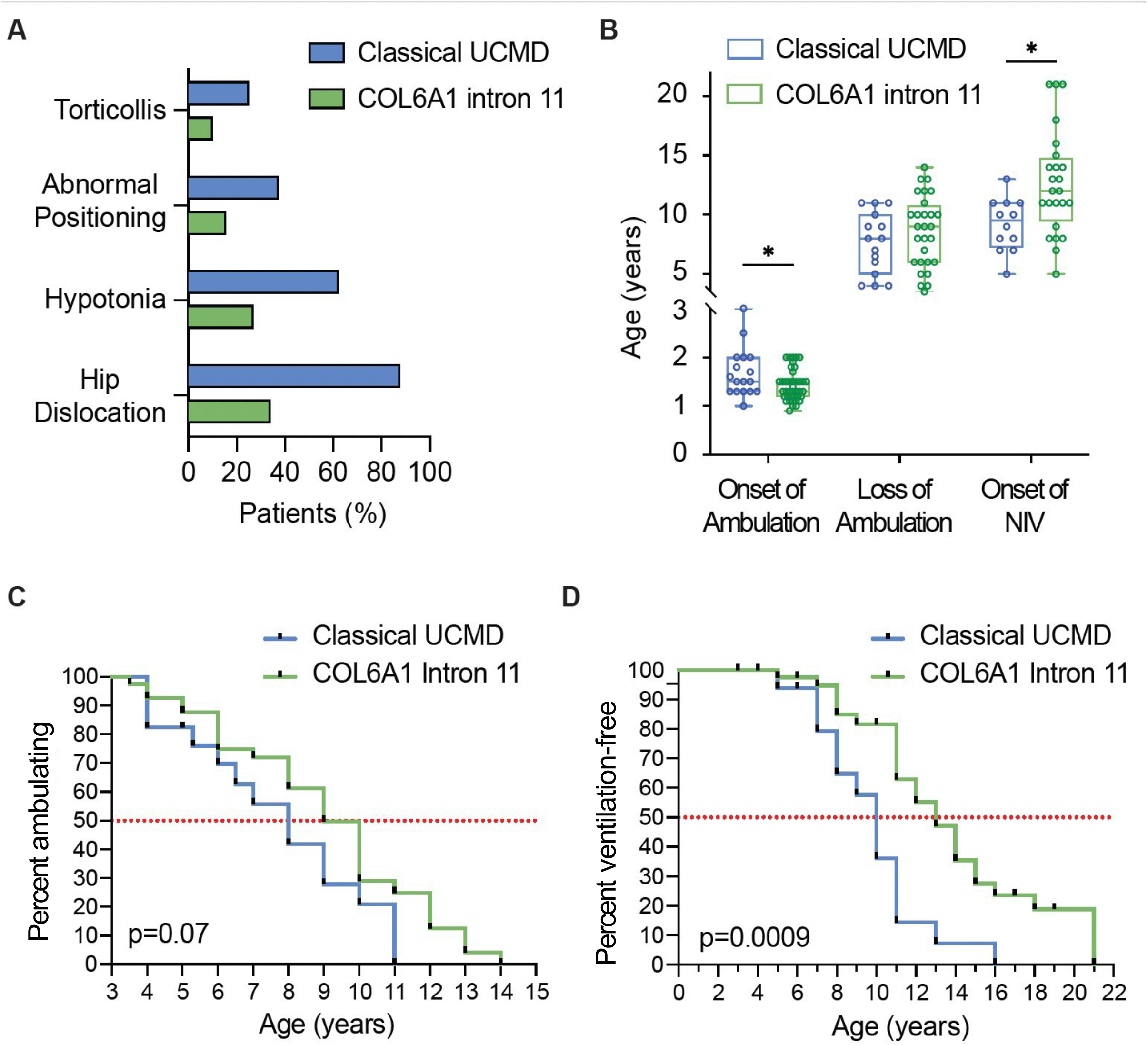
Clinical phenotype of patients heterozygous for the *COL6A1* c.930+189C>T variant versus patients with Classical UCMD (UCMD patients who do not have the *COL6A1* c.930+189C>T variant) **(A)** Bar graph demonstrating symptoms at birth in patients with the Classical UCMD phenotype (blue) versus the COL6A1 Intron 11 (green) phenotype **(B)** Box plot demonstrating distribution of ages at the time of major motor function and pulmonary function milestones. Onset of independent ambulation: (Classical UCMD phenotype, *n* = 17) (blue): box range 1.3 – 2.0; whiskers: 1.0 – 3.0. Onset of independent ambulation (COL6A1 Intron 11 phenotype, *n* = 40) (green): box range: 1.2 – 1.5; whiskers: 0.9 – 2.0 years. Loss of ambulation (Classical UCMD phenotype, *n* = 15) (blue): box range: 5.3 - 10.0; whiskers: 4.0 – 11.0. Loss of ambulation (COL6A1 Intron 11 phenotype, *n* = 28) (green): box range: 6-10.75; whiskers: 3.5 – 14. Onset on non-invasive ventilation (Classical UCMD phenotype, *n* = 12) (blue): box range: 7.3 – 11.0; whiskers: 5.0 – 13.0. Onset on non-invasive ventilation (COL6A1 Intron 11 phenotype, *n* = 24) (green): box range: 9.5 – 14.75; whiskers: 5 – 21. Asterisks indicate significance at the 0.05 level for Mann-Whitney U-tests. **(C)** Kaplan-Meier curve depicting independent ambulation in patients with Classical UCMD (blue) and patients with Intron 11 (green) (p=0.07) **(D)** Kaplan-Meier curve depicting ventilation-free status in patients with Classical UCMD (blue) and patients with Intron 11 (green) (p=0.0009)

The age at the time of the clinical evaluation ranges 3-38 years. Twenty-nine patients had lost independent ambulation (as defined by full-time wheelchair dependence) at the time of the clinical evaluation, with a mean age at loss of ambulation of 8.0 ± 3.0 years (Fig. 1B and C). Twenty-four patients had started non-invasive ventilation (NIV) in the form of bilevel positive airway pressure at the time of the clinical evaluation, at a mean age of 11.9 ± 4.4 years (Fig. 1B and D). Two patients in this cohort died: patient US9 during his 20s from probable *cor pulmonale*, in the setting of refusal to use non-invasive ventilation and patient CA3 during his 30s in the setting of respiratory failure and failure to thrive.

Joint contractures at the elbows, knees and wrists were assessed on clinical examination and categorised as ‘mild’: <45 degrees, ‘moderate’: 45 to <90 degrees and ‘severe’: >90 degrees. At the time of clinical evaluation, 7 (16%) patients had ‘mild’ joint contractures (ages 4-10 years), 16 (37%) patients had ‘moderate’ joint contractures (ages 3-16 years), and 20 (47%) patients had ‘severe’ joint contractures (ages 7-38 years) (Fig. 2A-E).

**Figure 2:** Joint contractures in heterozygous patients versus a patient with somatic mosaicism for *COL6A1* (c.930+189C>T) **(A)** Mild elbow contractures already noticeable in Patient US1; **(B)** Severe long finger flexor contractures in Patient IR1; **(C)** Severe wrist flexion and long finger flexor contractures in Patient US9; **(D)** Severe elbow, wrist flexion and long finger flexor contractures in Patient US5; **(E)** Severe elbow contracture (greater than 90 degrees in elbow flexion) in Patient US2. **(F)** Joint hyperlaxity of the elbow in Patient US16, who has somatic mosaicism for *COL6A1* (c.930+189C>T). *Please contact the corresponding author for access to these clinical images*.

Scoliosis requiring surgical repair was reported in 39% (15/38) patients with surgery performed in 10 patients, ranging in age at the time of surgery of 6-15 years. Three families declined scoliosis surgery, and two families opted to continue to observe the scoliosis at the time of the last assessment.

### Phenotype of patient with somatic mosaicism for the *COL6A1* c.930+189C>T variant

Patient US16 has a distinctly milder clinical phenotype. Her mother denied noting evidence of decreased fetal movement during the pregnancy. At the time of birth, hypotonia was noted, but there was no evidence of hip dislocation and/or hip dysplasia, abnormal positioning of hands and feet or torticollis. Patient US16 achieved independent ambulation without noted delay. At the time of her last clinical evaluation (first decade of life), she maintained the ability to ascend and descend stairs without holding onto the railing. She demonstrated the ability to ambulate independently with a Trendelenburg gait and was able to run with an exaggerated arm swing. Pulmonary function testing demonstrated a forced vital capacity (FVC) of 91% predicted upright and 79% predicted supine. There was evidence of only mild contractures of the long finger flexors and Achilles tendons and hyperlaxity of the elbow joints (Fig. 2F). There was no evidence of scoliosis.

### Phenotype of patients with UCMD phenotype due to causative variants in the *COL6* genes (other than the *COL6A1* c.930+189C>T variant)

Of those patients with findings documented at birth, 88% (15/17) had hip dislocation and/or hip dysplasia, 63% (10/16) had evidence of hypotonia, 38% (6/16) had abnormal positioning of hands and feet, and 25% (4/16) had torticollis (Fig. 1A). All patients had attained independent ambulation at a mean age of 1.6 ± 0.5 years (Fig. 1B). The age at the time of clinical evaluation ranges 5 to 29 years. Fifteen patients had lost independent ambulation (as defined by full-time wheelchair dependence) at the time of the clinical evaluation, with a mean age at loss of ambulation of 7.1 ± 2.6 years (Fig. 1B and C). Twelve patients had started non-invasive ventilation (NIV) in the form of bilevel positive airway pressure at the time of the clinical evaluation, at a mean age of 8.9 ± 2.3 years (Fig. 1B and D). One patient in this cohort died during the teenage years in the setting of respiratory failure and failure to thrive.

### Comparison of the phenotype of ‘classical UCMD’ and the phenotype of ‘COL6A1 Intron 11’

There was a statistically significant difference between ‘classical UCMD’ (due to causative variants in the *COL6* genes other than the *COL6A1* c.930+189C>T variant) and the ‘COL6A1 Intron 11’ phenotype (due to heterozygosity for the *COL6A1* c.930+189C>T variant) for the mean age at onset of independent ambulation (p = 0.04) while there was no statistically significant difference for the mean age at loss of ambulation (p = 0.32) (Fig. 1B). The difference between the mean age at onset of NIV between patients with classical UCMD and patients with the COL6A1 Intron 11 phenotype was statistically significant (p = 0.009) (Fig. 1B). Kaplan-Meier curves depicting the probability of independent ambulation (Fig. 1C) for patients with classical UCMD and COL6A1 Intron 11 did not demonstrate a statistically significant difference (p = 0.07) while Kaplan-Meier curves depicting ventilation-free probability (Fig. 1D) demonstrated a statistically significant difference (p = 0.0009).

### Muscle imaging

Muscle MRI (not available at all centres and challenging to perform in this cohort due to the need for non-invasive ventilation while in a supine position for the MRI) was performed in patient US10 at the NIH Clinical Center (Fig. 3A) and demonstrated abnormal signaling with a ‘central cloud’ pattern of abnormal signaling along the central fascia of the rectus femoris muscle and an ‘outside-in’ pattern of abnormal signaling along the periphery of the vastus lateralis, as is classically seen in patients with COL6-RD.^22–24^ Muscle MRI performed in patient US16 (who has somatic mosaicism for the *COL6A1* c.930+189C>T variant) (Fig. 3D) demonstrated mildly abnormal signaling in the vastus lateralis muscle.

**Figure 3:**
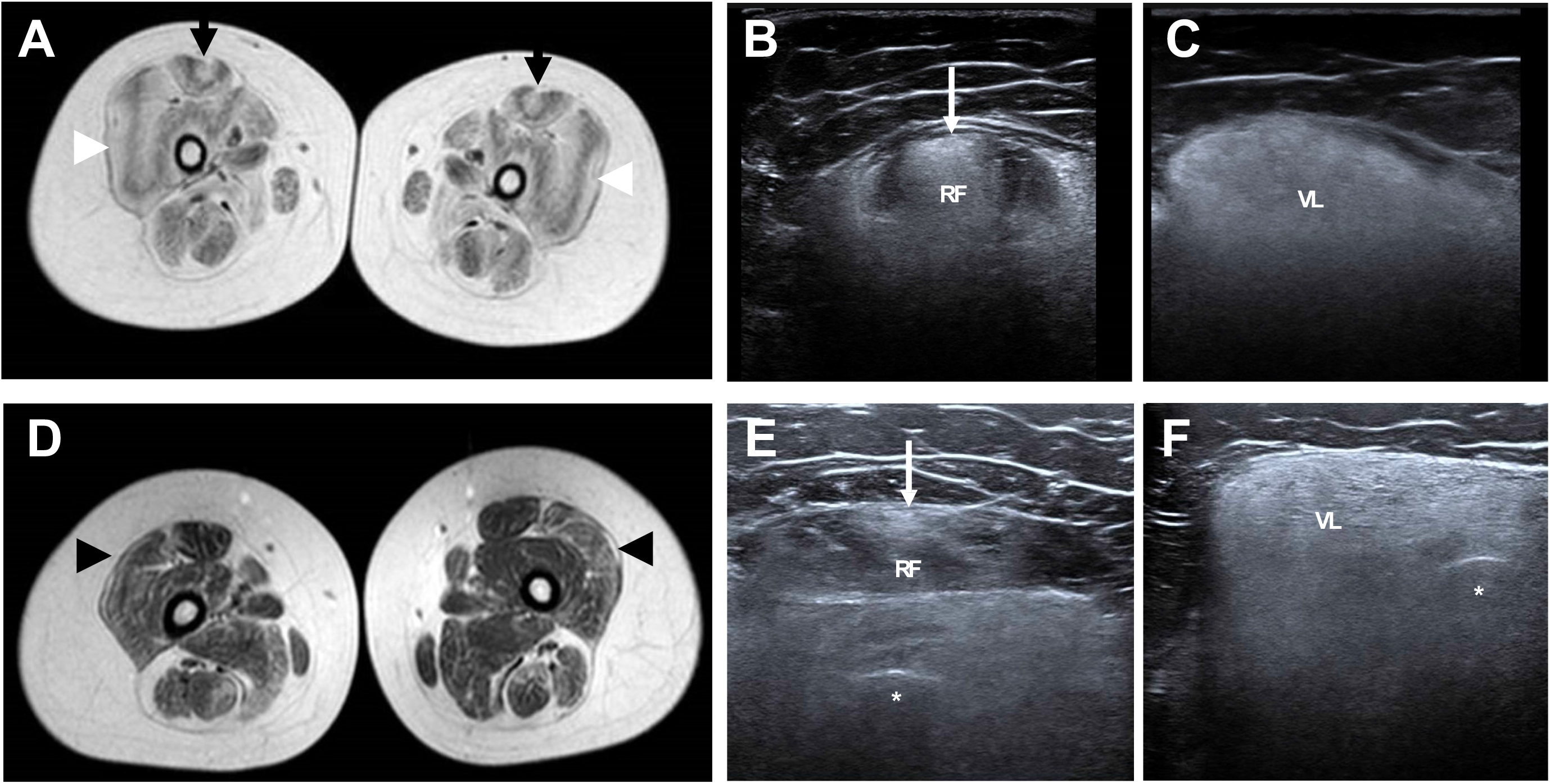
Muscle MRI and ultrasound in a heterozygous patient versus a patient with somatic mosaicism for *COL6A1* (c.930+189C>T) **(A)** Axial TI MRI of the upper leg in patient US10 demonstrating abnormal signaling consistent with a ‘central cloud’ pattern in the rectus femoris (black arrows) and an ‘outside in’ pattern in the vastus lateralis (white arrowheads); **(B)** Ultrasound of the rectus femoris (RF) muscle in patient US10 demonstrating abnormal signaling consistent with a ‘central cloud’ pattern (white arrow) with a loss of bone echogenicity; **(C)** Ultrasound of the vastus lateralis (VL) muscle in patient US10 demonstrating increased echogenicity with a loss of bone echogenicity**; (D)** Axial TI MRI of the upper leg in patient US16 demonstrating mildly abnormal signaling in the vastus lateralis muscle suggestive of a subtle ‘outside in’ pattern (black arrowheads); **(E)** Ultrasound of the rectus femoris (RF) muscle in patient US16 demonstrating an increase in echogenicity of the rectus femoris (RF) muscle consistent with a ‘central cloud’ pattern (white arrow) with bone echogenicity (asterisk) preserved**; (F)** Ultrasound of the vastus lateralis (VL) muscle in patient US16 demonstrating an increase in echogenicity of the vastus lateralis (VL) with bone echogenicity (asterisk) preserved.

Muscle ultrasound was performed in eight patients at the National Institutes of Health and demonstrated significantly increased echogenicity in a granular quality and a ‘central cloud’ pattern of increased echogenicity along the central fascia of the rectus femoris muscle and an ‘outside-in’ pattern of increased echogenicity along the outer region of the vastus lateralis, a classic muscle ultrasound pattern in patients with COL6-RD^23^ with loss of bone echogenicity, as appreciated in muscle ultrasound images (Fig. 3B and C). Muscle ultrasound performed in the mosaic patient US16 demonstrated increased echogenicity in a ‘central cloud’ pattern in the rectus femoris muscle with generally increased echogenicity in the vastus lateralis and maintenance of bone echogenicity (Fig. 3E and F), in contrast to patients heterozygous for *COL6A1* (c.930+189C>T) in whom bone echogenicity was lost due to the degree of increased echogenicity of muscles.

### Muscle immunofluorescence

Collagen VI immunofluorescence when performed on available muscle biopsy tissue demonstrated mislocalisation of collagen VI immunoreactivity, which was found to be accumulated in the interstitial space instead of colocalising with laminin at the basement membrane, as demonstrated in confocal microscopy images of the muscle biopsy of patient US1 (Fig. 4).

**Figure 4:**
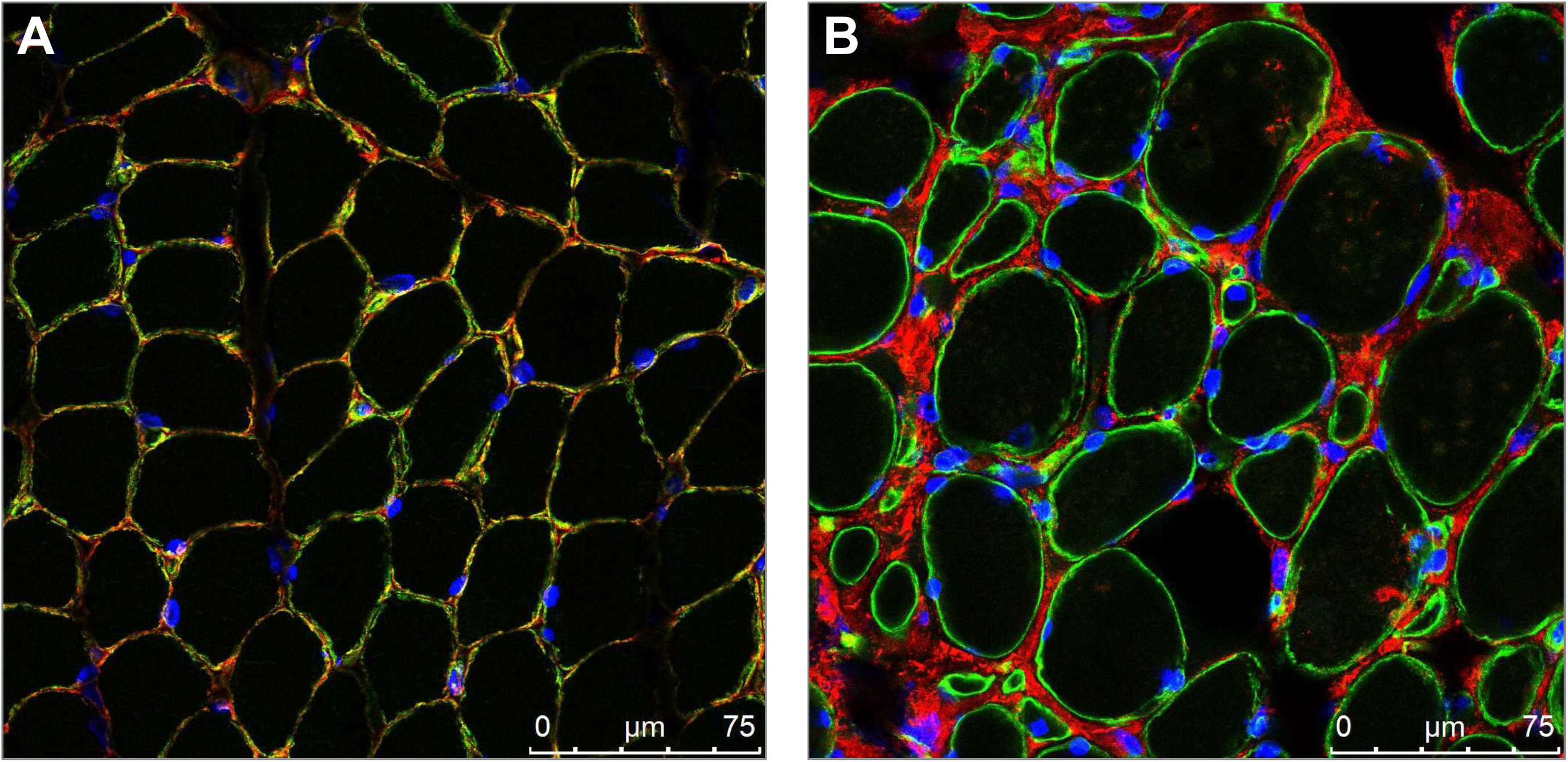
Muscle Immunofluorescence. Confocal imaging of muscle co-stained with collagen VI (red) and basement membrane marker laminin (green) along with nuclear stain DAPI (blue). **(A)** In control muscle, collagen VI is co-localised with laminin at the basement membrane. **(B)** In the muscle of patient US1 from a biopsy, collagen VI signal is observed in the interstitial space, indicative of collagen VI mislocalisation relative to the basement membrane. (magnification = 63X; scale bar = 75µm)

### Somatic mosaicism for *COL6A1* c.930+189C>T

Patient US16 was found to have somatic mosaicism for *COL6A1* c.930+189C>T based on Sanger sequencing of genomic DNA performed in various tissues (Fig. 5A). The degree of mosaicism, calculated from the fractional abundance (or percent) of the variant (‘T’) allele as measured by digital droplet PCR, was determined to be ~20%, meaning that approximately 40% of cells in this individual harbour the c.930+189C>T variant (Fig. 5B). Tissues tested were derived from the endoderm (bladder epithelium in the urine sample), the mesoderm (blood cells, dermis of the skin biopsy sample), or the ectoderm (epidermis of the skin biopsy sample, buccal epithelium in the saliva sample) (Fig. 5B). Consistent with this finding, expression levels of *COL6A1* transcripts including the pseudoexon as assessed in a fresh skin biopsy sample, and in cultured dermal fibroblasts, were lower in US16 compared to samples obtained in patients heterozygous for *COL6A1* c.930+189C>T (Fig. 5C). By quantitative PCR performed in fresh skin biopsies, we determined that the *COL6A1* transcripts with inclusion of the pseudoexon were 7.2-fold lower in the patient with somatic mosaicism for *COL6A1* c.930+189C>T (US16) compared to a patient heterozygous for *COL6A1* c.930+189C>T (US5) (Fig. 5D), although additional samples would be needed to determine statistical significance.

**Figure 5:**
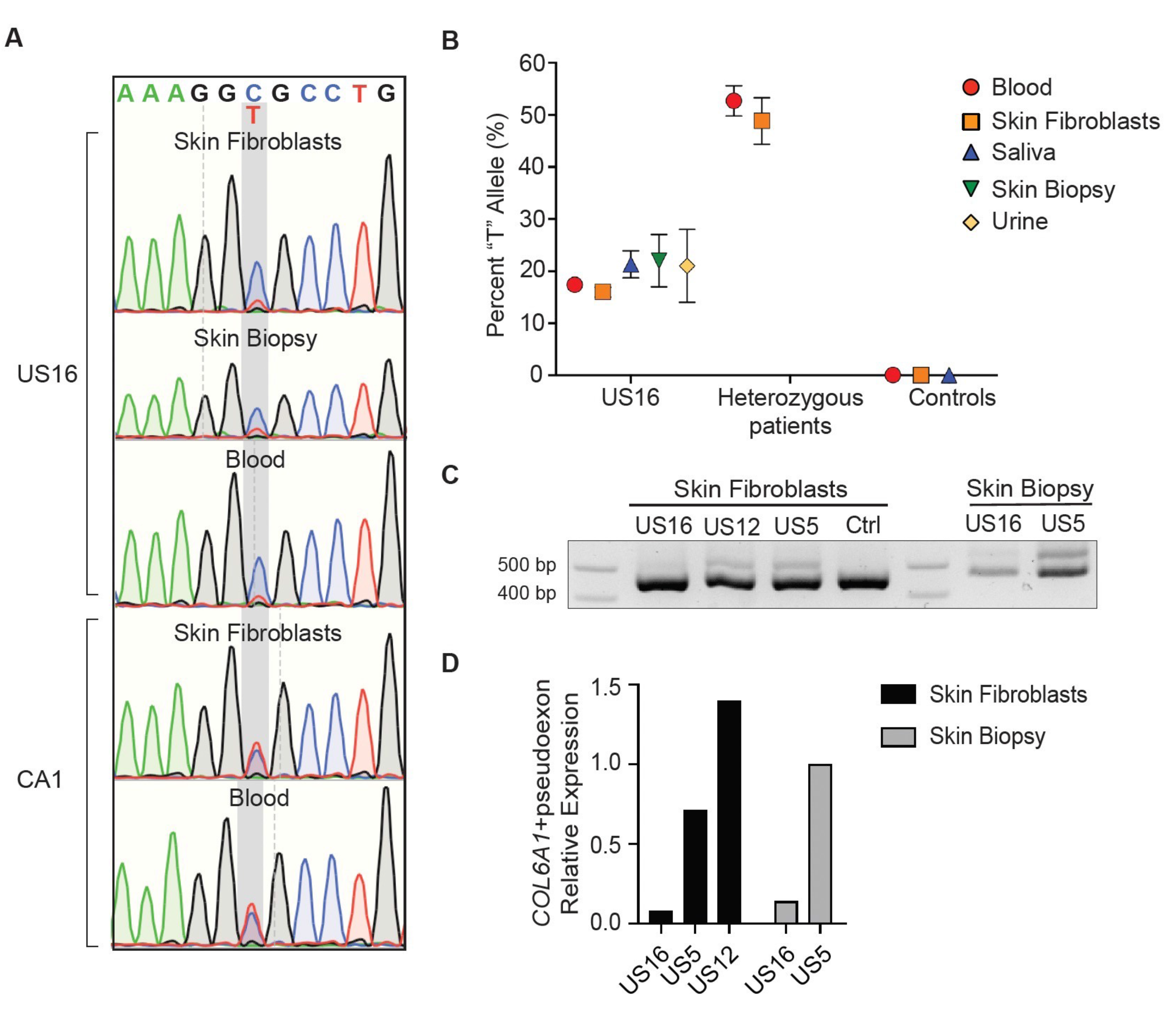
Somatic mosaicism for *COL6A1* (c.930+189C>T) **(A)** Genomic DNA sequencing chromatograms at the *COL6A1* (c.930+189C>T) locus showing comparable peak heights for the cytosine and thymine alleles in a patient heterozygous for *COL6A1* (c.930+189C>T) (CA1), but a higher cytosine peak height compared to thymine in the patient mosaic for *COL6A1* (c.930+189C>T) (US16), in various tissue samples (skin fibroblasts, skin biopsy, and blood). **(B)** Determination of the degree of mosaicism was achieved by droplet digital PCR quantification using a genotyping probe assay and genomic DNA as input. Graph shows the fractional abundance of the thymine (“T”) allele, calculated as the ratio of “T” concentration (copies/uL) over total (“T” + “C”) concentration (copies/uL). Error bars represent the Poisson confidence interval. [The heterozygous patients were patient HK1 (blood) and patient US12 (skin fibroblasts).] (**C)** RNA isolated from skin biopsies or from skin-derived primary fibroblasts was reverse-transcribed and amplified with primers spanning *COL6A1* exons 10 to 20. In patient US16 [mosaic for *COL6A1* (c.930+189C>T)] the upper band (transcripts with pseudoexon) appears fainter than in US12 and US5 [heterozygous for *COL6A1* (c.930+189C>T)]. **(D)** Relative expression of *COL6A1* with pseudoexon transcripts normalised to total *COL6A1* levels in skin fibroblasts and skin biopsy of the patient mosaic for *COL6A1* (c.930+189C>T) (US16), compared to two patients heterozygous for *COL6A1* (c.930+189C>T) (US5 and US12). A fresh skin biopsy was not available for patient US12.

## Discussion

The COL6-related dystrophies typically manifest symptoms at birth and thus are at their core congenital muscular dystrophies. The COL6-RD subtype Ullrich congenital muscular dystrophy (UCMD) is characterised by prominent symptoms at the time of birth including significant hypotonia, proximal joint contractures, distal hyperlaxity, abnormal positioning of the hands and feet, prominent calcanei, hip dislocation(s), torticollis and kyphoscoliosis.^1,6–9^ Here we report an international cohort of 44 patients all habouring a *de novo COL6A1* c.930+189C>T deep intronic pseudoexon-inducing variant, and we delineate their presentation and natural history in comparison to patients with classical UCMD.

A hallmark of this *COL6A1* c.930+189C>T-specific cohort is that all patients who are heterozygous for this causative variant (N=43) demonstrate a paucity of congenital symptoms, followed by an apparent accelerated progression of symptoms, ultimately demonstrating a phenotype consistent with the well-defined phenotype of UCMD (as distinguished from other COL6-RD phenotypes),^2,12,14,25^ what we will refer to as ‘classical UCMD.’ In fact, only approximately one third or less of the patients heterozygous for *COL6A1* c.930+189C>T had evidence of any symptoms at birth. Furthermore, with a mean age of 1.4 years at the onset of independent ambulation, most patients in this *COL6A1* c.930+189C>T-specific cohort did not present to a neurologist until the time of delayed independent ambulation or afterwards, when evidence of difficulty arising from the floor, or frequent falls were noted. In contrast, in a comparison cohort of patients with the UCMD phenotype who do not harbour the *COL6A1* c.930+189C>T variant (N=17), symptoms at birth were present in up to 88%, thus typically prompting a neurological evaluation congenitally.

The mean age at loss of independent ambulation in this *COL6A1* c.930+189C>T-specific cohort is 8.0 ± 3.0 years, which is not statistically different from our comparison cohort of patients with classical UCMD (non *COL6A1* c.930+189C>T UCMD; N=17) of 7.1 ± 2.6 years (Fig. 6). The mean age at the time of initiation of non-invasive ventilation (NIV) in patients heterozygous for this *COL6A1* deep intronic variant is 11.9 ± 4.4 years. Of note, while this mean age is statistically different from the mean age at the time of NIV initiation of 8.9 ± 2.3 in our comparison cohort of classical UCMD, it is similar to the data in the largest international natural history study of pulmonary function in patients with UCMD (N=75) in which the mean age at the time of NIV initiation was 11.3 ± 4.0 years^2^ (Fig. 6). Given differences in practice among centres internationally relating to frequency of pulmonary function testing and polysomnogram assessments as well as thresholds for starting NIV, it likely would be more accurate to compare the international *COL6A1* c.930+189C>T-specific cohort (N=43) to the international classical UCMD cohort (N=75) than to the comparison cohort of classical UCMD (N=17) evaluated at one center where the initiation of NIV tends to be more proactive.

**Figure 6:**
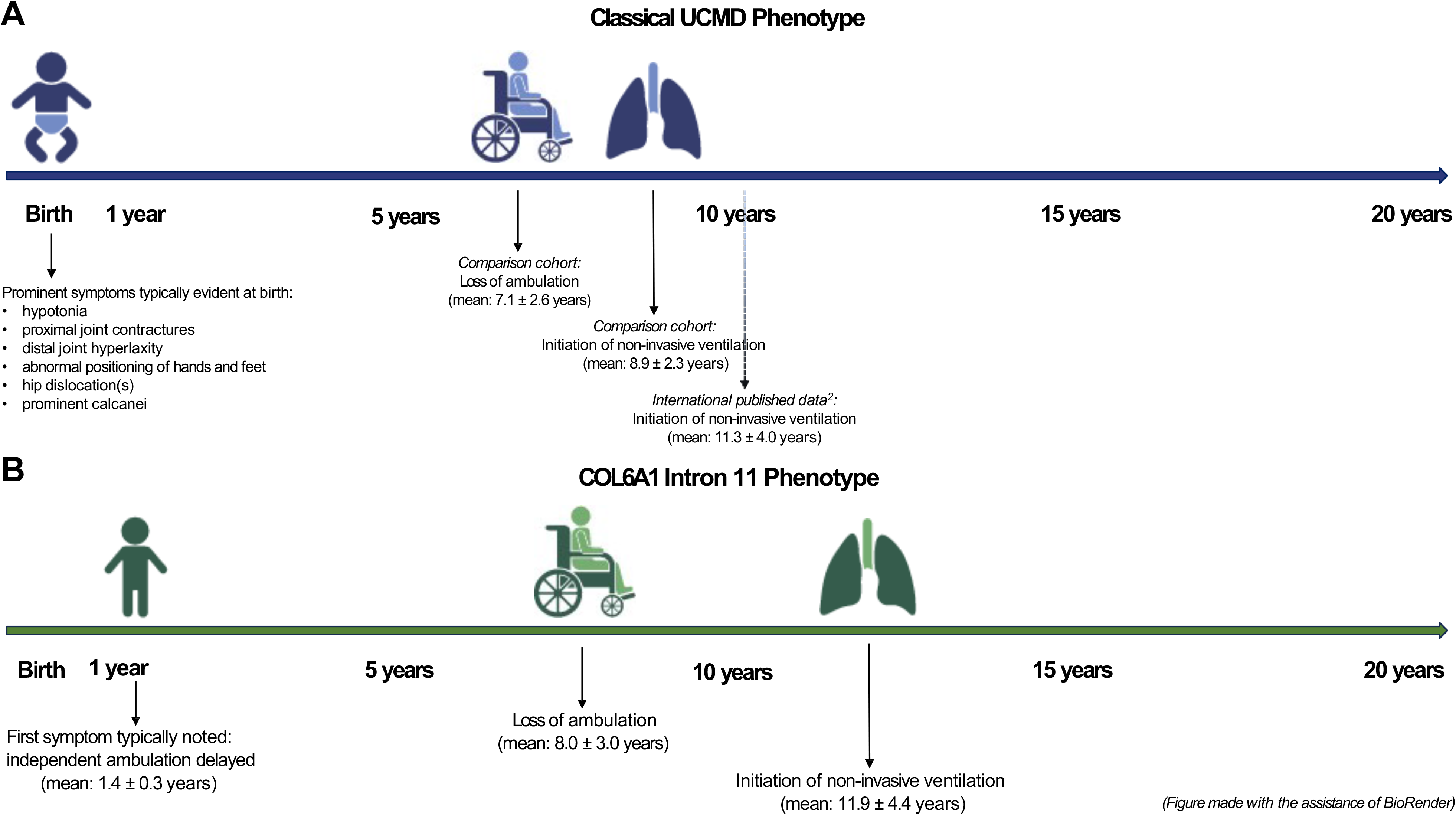
Comparison of the COL6A1 Intron 11 Phenotype and the Classical UCMD Phenotype. **(A)** Schematic of the natural history of the motor and pulmonary function in patients with the classical UCMD phenotype in the comparison cohort (N=17). Natural history of pulmonary function data from the largest published international natural history study of patients included (N=75)^2^ (dotted line). **(B)** Schematic of the natural history of motor and pulmonary function in this cohort of patients who are heterozygous for *COL6A1* (c.930+189C>T) (N=43)

Overall, what distinguishes this *COL6A1* c.930+189C>T-specific cohort phenotype from the classical UCMD phenotype is a delayed and then accelerated rate of progression of symptoms in comparison to classical UCMD, in which striking symptoms are evident at the time of birth (Fig. 6). All patients who are heterozygous for this causative *COL6A1* intron 11 variant (not with somatic mosaicism) ultimately arrive at a highly consistent clinical severity of motor and likely also pulmonary function (depending on the comparator cohort), characterised by loss of ambulation and respiratory insufficiency at approximately the same age as classical UCMD, despite manifesting first symptoms at a mean age of 1.4 ± 0.3 years (Fig. 6). Given this convergence of phenotypic features in patients in this *COL6A1* c.930+189C>T-specific cohort and patients with classical UCMD, outcome measures validated in patients with the classical UCMD phenotype could be used for patients with the *COL6A1* Intron 11 phenotype.^26,27^

In COL6-RDs, respiratory insufficiency is largely due to disproportionate weakness of the diaphragm.^2,28^ In this particular cohort of patients with *de novo COL6A1* c.930+189C>T, it is possible that the severity of joint contractures may contribute to further decreasing the compliance of the chest wall, thus likely further exacerbating patients’ respiratory insufficiency. It is important to note that the two patients in this cohort who passed away had evidence of severe respiratory insufficiency as well as severe joint contractures (greater than 90 degrees). Patient US9 had been prescribed NIV in the form of bilevel airway pressure during the teenage years but refused to use NIV and passed away in his 20s (with last recorded FVC of 21% three years prior to his death). Patient CA3 had evidence of respiratory failure (with last recorded FVC of 8%) and failure to thrive prior to passing away in his 30s.

Patients heterozygous for *COL6A1* c.930+189C>T have muscle imaging findings including muscle ultrasound and muscle MRI findings which are consistent with those findings described in association with COL6-RDs.^22–24^ Thus, muscle imaging remains a very helpful tool in supporting efforts to identify a causative variant in the *COL6* genes in the setting of a clinical suspicion of COL6-RD. Furthermore, muscle immunohistochemistry studies in patients harbouring *COL6A1* c.930+189C>T demonstrate mislocalised collagen VI expression, as classically seen in COL6-RD due to dominant mutational mechanisms.^5^ Thus, in the setting of clinical examination findings and muscle imaging findings suggestive of COL6-RD as well as in the setting of evidence of mislocalisation of collagen VI on muscle immunohistochemistry, it is essential to consider the diagnostic possibility of COL6-RD due to *COL6A1* c.930+189C>T and to ensure that genetic testing laboratories adequately assess for the presence of this deep intronic variant.

The exact pathomechanisms by which the *COL6A1* c.930+189C>T causative variant results in a severe phenotype of UCMD with an accelerated progression of symptoms in comparison to classical UCMD were partially elucidated. Strikingly, we found that approximately 25% of total *COL6A1* transcripts include the 72-nt pseudoexon in muscle tissues, as opposed to the expected 50%, due to the “leakiness” of the c.930+189C>T variant,^19,20^ which could explain the initial delay in the presentation of symptoms. The mutant collagen α1(VI) protein produced includes a stretch of 24-amino acid residues that disrupts the Gly-X-Y repeat in its amino-terminus,^20^ prior to the cysteine residue involved in dimerisation, a ‘hot-spot’ where dominant-negative *COL6* variants are known to allow mutant chains’ assembly into tetramers. It can thus be assumed that the mutant collagen α1(VI) protein containing the pseudoexon-encoding sequence exerts a dominant-negative effect by assembling into tetramers and consequently disrupting polymerisation of collagen VI tetramers. In fact, costaining with a mutation-specific antibody and a collagen α3(VI)-N-terminus-specific antibody showed a broad overlap in immunofluorescence microscopy in patient muscle.^29^ However, when patient fibroblast cell culture supernatants were studied by composite agarose/polyacrylamide gel electrophoresis and immunoblotting, it was found that the mutant collagen α1(VI) was secreted as single chains.^29^ Interestingly, like in the supernatant from healthy control fibroblasts, wild-type collagen VI tetramers were found in the supernatant of patient fibroblast cultures. Indeed, in negative stain electron micrographs of immunogold-labelled supernatants of patient fibroblasts, collagen VI microfibrils with the typical spacing of the globular beads were detected with a gold-labeled collagen α3(VI)-N-terminus antibody. In contrast, the gold-labelled mutation-specific antibody bound to protein aggregates and sometimes decorated collagen VI microfibrils.^29^ The two effects of aggregated mutant collagen α1(VI) chains and the decoration of collagen VI microfibrils are not mutually exclusive and could be cumulative, which may explain the acceleration of clinical symptoms once the disease starts.

The strikingly milder clinical phenotype observed in patient (US16) who has somatic mosaicism for the *COL6A1* c.930+189C>T variant, for whom expression of pseudoexon transcripts was 7.2-fold lower compared to the heterozygous patient US5 (as assessed in respective skin biopsies), highlights how in principle a reduction in the abundance of the pseudoexon would translate to an amelioration of clinical symptoms. In particular, the observation that patient US16 continues to ascend and descend stairs without use of the railing suggests a phenotype consistent with Bethlem muscular dystrophy.^25^ In keeping with this patient’s milder motor phenotype is her mildly affected pulmonary function, with an FVC of 91% upright and 79% supine. Thus, this patient represents an *in vivo* scenario of our previously published *in vitro* rescue of the pseudoexon insertion with splice-modulating antisense oligomers which effectively decreased the levels of pseudoexon transcripts by ~7-fold (for PMO-PEX1, at the highest concentration tested, in cultured fibroblasts) and consequently levels of the aberrant protein product. ^20^

Taken together, given the clinical severity of UCMD described in this large international cohort of patients harbouring *COL6A1* c.930+189C>T, the recognition that this variant is one of the most common recurrent causative variants in COL6-RDs and the promise of therapeutic rescue as demonstrated by our pseudoexon skipping/splice-modulating *in vitro* work,^20,30,31^ it is imperative that patients harbouring this deep intronic variant are clinically recognised and diagnosed. To this end, it is necessary for next generation sequencing panels to include an algorithm to ensure that intron 11 of *COL6A1* is captured, including libraries built for high throughput which would capture this variant. Moreover, we suggest designating the *COL6A1* c.930+189C>T-specific phenotype described here as a ‘COL6A1 Intron 11’ phenotype, given the need to distinguish this phenotype characterised by a delayed onset of clinical symptoms followed by an accelerated progression of symptoms from other patients with COL6-RD for the purpose of natural history studies in preparation for future clinical trials. Distinguishing this so-called ‘COL6A1 Intron 11’ phenotype is essential from the perspective of inclusion criteria for clinical trial stratification, including for non-variant-specific therapeutic approaches for COL6-RDs, such as therapeutic approaches targeting TGFβ, fibrosis and apoptosis. Our characterization of this *COL6A1* c.930+189C>T-specific cohort including the comparative retrospective natural history of patients with COL6A1 Intron 11 to patients with classical UCMD contributes to the clinical trial readiness of the COL6A1 Intron 11 patient population, who are facing the promise of the *COL6A1* c.930+189C>T variant-specific splice-modulating therapeutic approach in development.^20,30,31^

## Acknowledgements

We especially thank the patients and their families whose participation made this study possible.

Figure 6 was made with the assistance of Biorender.com.

## Supplementary Material

Supplementary material is available online.

## Data Availability

All deidentified data are available upon request from the corresponding author.

---

**Supplementary Table 1: Core phenotypic features of patients with UCMD due to causative variants in the COL6 genes (other than the *COL6A1* c.930+189C>T variant)**

^†^Died (related to respiratory insufficiency)

NA = not available

