## Supplementary Table 1 for "The recurrent deep intronic pseudoexon-inducing variant *COL6A1* c.930+189C>T results in a consistently severe phenotype of COL6-related dystrophy: Towards clinical trial readiness for splice-modulating therapy"

**Supplementary Table 1: Core phenotypic features of patients with UCMD due to variants in the COL6 genes (other than the *COL6A1* c.930+189C>T variant)**

| **Patient Identifier** | **Causative variants in the COL6 genes *(COL6A1*, *COL6A2* or *COL6A3*)** | **Sex** | **Race/Ethnicity** | **Hip dislocation or hip dysplasia at birth** | **Hypotonia at birth** | **Abnormal position of hands/feet at birth** | **Torticollis at birth** |
| --- | --- | --- | --- | --- | --- | --- | --- |
| P1 | *COL6A3*: c.6181C>T, p.Arg2061*; c.6931G>A, p.Gly2311Arg | F | Caucasian/American | Y | Y | N | N |
| P2 | *COL6A3*: c.6210+1G>A | M | Caucasian/American | Y | Y | N | N |
| P3 | *COL6A1*: c.859G>A, p.Gly287Arg | M | Caucasian/American | N | N | Y | N |
| P4† | *COL6A1*: c.850G>A, p.Gly284Arg | F | Caucasian/American | Y | NA | NA | NA |
| P5 | *COL6A3*: c.6309+1G>T | F | Caucasian/American | Y | Y | Y | Y |
| P6 | *COL6A2*: c.801+2_801+3insTTT | M | Caucasian/American | Y | N | Y | N |
| P7 | *COL6A2*: c.2386A>T, p.Lys796*; c.856-3C>G | F | Asian/Korean | Y | Y | Y | Y |
| P8 | *COL6A1*: c.868G>A, p.Gly290Arg | M | Caucasian/American | Y | Y | N | N |
| P9 | *COL6A1*: c.860G>A, p.Gly287Glu | F | Caucasian/American | Y | N | N | N |
| P10 | *COL6A2:* c.855+2T>G | F | Caucasian/American | Y | Y | Y | Y |
| P11 | *COL6A1*: c.957+2T>C | F | Caucasian/American | Y | Y | Y | N |
| P12 | *COL6A1*: c.850G>A, p.Gly284Arg | F | Caucasian/American | Y | N | N | Y |
| P13 | *COL6A1*: c.904G>C, p.Gly302Arg | F | Caucasian/American | Y | N | N | N |
| P14 | *COL6A2*: c.855+1G>T | M | Caucasian/Polish | N | Y | N | N |
| P15 | *COL6A1*: c.957+1G>T, p.Gly311_Asn319del | F | Hispanic/Mexican | Y | Y | N | N |
| P16 | *COL6A3*: g.238269008_238273480delins238264873_238265197inv | F | Caucasian/Mixed European | Y | Y | Y | N |
| P17 | *COL6A3*: c.6212_6309+29del | F | Caucasian/American | Y | N | N | N |

^†^Died (due to respiratory failure and failure to thrive)

NA = not available
